# Effect of Adapted Ergometer Setup and Rowing Speed on Lower Extremity Loading in People with and without Spinal Cord Injury

**DOI:** 10.1101/2021.02.08.21251109

**Authors:** Ying Fang, Karen L. Troy

## Abstract

**Introduction:** Functional electrical stimulation assisted rowing (FES-rowing) on an adapted ergometer is used in spinal cord injury (SCI) rehabilitation. A primary goal is to mechanically load the lower extremity to prevent disuse osteoporosis. Recent studies reported the small foot reaction force in FES-rowing was not sufficient to prevent bone loss.

**Objective:** This study aims to investigate the effect of ergometer setup and rowing speed on lower extremity loading in able-bodied and SCI individuals.

**Design:** Twenty able-bodied novice rowers and one experienced SCI rower rowed on an adapted ergometer with different speeds and setups. Motion capture system and force sensors were used to calculate forces and moments at the knee.

**Main Outcome Measures:** Foot reaction force and knee moment for all participants, and tibiofemoral force of the SCI rower.

**Results:** Peak foot reaction forces of able-bodied rowers ranged from 0.28 – 0.45 body weights (BW), which was less than half the force in normal rowing. A fast rowing speed, forward seat position, and large knee RoM were associated with higher foot force and knee moment during able-bodied rowing. The SCI subject had the greatest foot reaction force (0.40 BW) when rowing with small knee RoM at a rear seat position and the highest tibiofemoral force (2.23 BW) with large knee RoM at a rear seat position.

**Conclusion:** Ergometer setup and speed can double the force generation at the foot during both able-bodied rowing and FES-rowing. Rowing forms (range of motion and speed) that resulted in the greatest foot reaction force were different for able-bodied rowers and SCI rowers, indicating a trade-off between motion and force generation in FES-rowing that warrants further investigation with more SCI rowers. Clinicians and physical therapist should be aware that ergometer setups can be easily adjusted to modify rowing forms and loading patterns of users with SCI.

## Introduction

People with spinal cord injury (SCI) experience severe bone loss in their lower limbs, leading to a 40% long-term fracture risk^1^. Physical activity could improve bone health in individuals with SCI by reloading their lower extremities^2–4^. A dose-response relationship has been observed with greater loading to the musculoskeletal system leading to less bone loss, for activities such as functional electrical stimulation assisted (FES) cycling and electrical stimulation-induced standing^5–7^.

FES-rowing is one exercise intervention that is growing in popularity in SCI rehabilitation^8^. Previous studies reported inconsistent bone responses after training with FES-rowing^9–11^ and a wide range of foot reaction forces generated by SCI rowers (0.22 to 0.67 body weight (BW))^12–14^. These variable results, combined with observations that bone adapts to its mechanical loading environment, suggest that foot loading influences subsequent changes in lower extremity bone in FES-rowing. Furthermore, it suggests that people who benefit from the exercise may row differently from those who do not. However, little is known about the potential factors that may influence force generation in FES-rowing.

In able-bodied rowing, both rowing speed and rowing experience affect the magnitude of foot reaction force, and consequently, joint loading. Elite rowers generate significantly higher foot reaction force and knee moment than novice or non-rowers^15^. Rowing speed influences force production in novice rowers, but not experienced rowers^15,16^. The design and setup of the ergometer also affect force production^17,18^. For example, increasing the height of the foot stretcher significantly reduced foot force and ankle, knee, and hip moments among elite rowers^19^. There are three major changes to a normal ergometer to make it suitable for FES-rowing: 1) two stoppers are added on the seat rail to limit the anterior-posterior movement of the seat, which prevents knee hyperextension; 2) a seat backrest is added, and the rower wears a seatbelt for stabilization, so there is no trunk flexion/extension; 3) a knee stabilizer is placed between the legs during rowing to prevent the knees from separating apart. These changes add additional constraints to the activity and may alter rowing biomechanics. They also allow more adjustments on the ergometer, which could affect force production by the SCI users.

Considering the recent finding that foot loading was lower than expected in FES-rowing, the purpose of the study was to investigate the effect of rowing speed, knee range of motion (RoM), and seat position on lower extremity loading in a group of able-bodied adults and a single individual with SCI. Specifically, we focused on foot reaction force and knee extension moment in all participants and evaluated tibiofemoral in the SCI subject. We hypothesized that rowing speed, knee RoM, and seat position would all affect the foot reaction force and knee moment produced by able-bodied rowers and the SCI rower, as well as affect peak tibiofemoral force of the SCI rower.

## Methods

### Subjects

Ten male and ten female able-bodied adults (age: 26.5 ± 3.8 years, mass: 70.0 ± 14.8 kg, height: 1.7 ± 0.1 m) with no rowing experience or lower extremity injuries within the past six months, and one male with complete SCI in his 20’s (mass: 75.0 kg, height: 1.7 m, FES-rowing experience: > 5 years) participated in this study. Each participant read and signed an informed consent document approved by the institutional review board before testing. Power analysis indicated that a sample size of 14 was enough to detect a within-subject difference in peak foot reaction force of 30 ± 20 N with 80% power for conditions with different setups (α = 0.05).

### Instrumentation

A ten-camera motion capture system (100 Hz; Vicon Motion Analysis Inc., UK) was used to collect 3D kinematic data. Reflective markers were placed bilaterally on the acromion processes, anterior superior iliac spines, great trochanters, medial and lateral epicondyles of the knee, medial and lateral malleoli, and the middle toes. Two markers were adhered to the seatback to represent the posterior superior iliac spines, which were blocked by the seatback during rowing. The above markers were used to define the trunk, pelvis, thighs, lower legs, and feet of the subject.

An adapted rowing ergometer (Concept2, model D, Morrisville, VT, USA), commonly used in clinical settings for FES-rowing, was instrumented with several force sensors and used for all tests. A 6 degree of freedom (dof) force sensor (1000 Hz; MC3A-1000lb, AMTI, Watertown, MA, USA) was mounted under the right foot stretcher of the ergometer to record the foot reaction forces and moments. A dummy block with the same geometry and weight of the sensor was mounted under the left foot stretcher to ensure both sides were symmetrical. During FES-rowing, the SCI participant brought his own electrical stimulator (Odstock, Salisbury, UK) along with a hand switch to control the stimulation. The 4 channel stimulator generated electrical signals (No ramp, pulse width: 450 μs, frequency: 40 Hz) to contract the knee extensors and flexors: pressing the switch activates the quadriceps and releasing the switch activates the hamstrings^20^. Signals from the force sensor were amplified and filtered using a signal conditioner (GEN 5, AMTI, Watertown, MA, USA) and recorded within the VICON system and its Nexus software simultaneously with the kinematic data.

### Experimental protocol

Upon arrival to the lab, able-bodied participants were instructed to row on the adapted ergometer and were given sufficient time to practice until a smooth movement was achieved. Then, individuals sat on the seat with both feet resting on the foot stretchers, and a researcher determined the location of the front and rear stoppers based on the participant’s knee angle. Specifically, the front stopper was adjusted such that the minimum knee angle was 45°, 70°, or 95°; the rear stopper was adjusted such that the maximum knee angle was 115°, 135°, 140°, or 165°. In this way, the seat position and knee RoM could be independently adjusted (Table 1). Participants rowed for 90 seconds at each of the 12 conditions that include three speeds (25, 35, and 40 SPM with a forward seat position and knee RoM of 70° and 90°), three knee RoM (70°, 90°, and 120° with a forward seat position at speed of 25 SPM and 35 SPM), and three seat positions (forward, middle, and rear, with knee RoM of 70° at speed of 25 SPM and 35 SPM).

**Table 1.** Required rowing speed and form for each of the 12 conditions tested in able-bodied participants. Shaded area indicates conditions that were tested on the participant with spinal cord injury (SCI). RoM: range of motion. SPM: stroke per minute.

| Condition | Speed (SPM) | Knee RoM (°) | Knee Angle Range (°) | Seat Position |
| --- | --- | --- | --- | --- |
| 1 | 40 | 70 | 45 - 115 | Front |
| 2 | 40 | 90 | 45 - 135 | Front |
| 3 | 25 | 70 | 45 - 115 | Front |
| 4 | 35 | 70 | 45 - 115 | Front |
| 5 | 25 | 90 | 45 - 135 | Front |
| 6 | 35 | 90 | 45 - 135 | Front |
| 7 | 25 | 120 | 45 - 165 | Front |
| 8 | 35 | 120 | 45 - 165 | Front |
| 9 | 25 | 70 | 70 - 140 | Middle |
| 10 | 35 | 70 | 70 - 140 | Middle |
| 11 | 25 | 70 | 95 - 165 | Rear |
| 12 | 35 | 70 | 95 - 165 | Rear |

The FES-rowing test followed similar experimental procedures. The individual with SCI applied the electrodes and set up the stimulator himself, and the researcher determined the location of stoppers. To avoid fatigue, this participant tested 7 rowing conditions: a self-selected style and 6 out of the 12 conditions that were tested in able-bodied rowers. The six subsets were selected because they yielded the largest foot reaction forces in able-bodied subjects. For both able-bodied and FES-rowing, resistance on the ergometer was set at level 3, which is commonly used for FES-rowing based on previous interviews with 4 SCI rowers. Rowing speed was controlled by the real-time feedback on the monitor of the ergometer. Data were collected for 30 s starting from 30 s into the bout of rowing for each condition.

### Data Analyses

The raw kinematics and force data were filtered using a low-pass fourth-order Butterworth filter at a cutoff frequency of 1 Hz. Joint angles were calculated based on joint centers and coordinate systems that were defined using ISB recommendations^21^. Specifically, knee angle was defined as 180° when the knee was fully extended. Joint moments and forces were calculated with inverse dynamics using a recursive Newton-Euler approach^22^ and averaged for at least five rowing cycles under each condition. The beginning of each cycle was defined when the seat was at the front-most position. We used customized code to perform all data analyses in Matlab (MathWorks, Natick, MA, USA). Peak foot reaction forces and knee extension moments were the primary outcomes of interest.

Tibiofemoral force of the SCI rower was calculated using OpenSim 3.3^23^. First, we developed a musculoskeletal model of the SCI participant by scaling a full-body generic model^24^ using experimental markers, and the shoulder, elbow, and wrist joints of the model were fixed. Mass and inertial properties of each segment were initially scaled based on the participant’s mass^25^, and then updated according to the anthropometric data specific to the SCI population^26^. Marker trajectories of each trial were applied to the scaled model to replicate experimentally measured kinematics. We assumed both legs generated symmetrical force and used the measured force under the right foot to estimate the force under the left foot. We also assumed all forces beyond the hip (e.g., hand force, seat force, etc) to be a resultant force applying at the pelvis (pelvis force), which was derived from dynamic analysis. We used static optimization to estimate muscle forces. Based on the electrode location, the quadriceps corresponded to rectus femoris, vastus lateralis, and vastus medialis, and the hamstrings corresponded to biceps femoris long head and semitendinosus in the model. The remaining muscles were enabled in the model with a fixed activation level of 0.01 to account for passive stiffness. Using the joint reaction analyses tool, we resolved tibiofemoral force considering muscle forces, external forces, and joint kinematics. Here we report the knee compressive tibiofemoral force.

### Statistical Analyses

For able-bodied rowing, repeated measures analyses of variance was used to examine the influence of each of the three rowing factors: rowing speed, knee RoM, and seat position, on the peak foot reaction force and peak knee extension moment. If a main effect was significant, post hoc analysis was performed using pairwise t-tests with Bonferroni adjustments. All analyses were performed in SPSS (IBM SPSS Statistics 22, Chicago, IL, USA) with an alpha level of 0.05.

## Results

Peak foot reaction force ranged from 0.28 to 0.45 BW across the 12 conditions in able-bodied rowers, with no significant difference between males and females. Peak foot reaction force and peak compressive tibiofemoral force ranged from 0.26 – 0.40 BW and 0.04 – 2.23 BW, respectively, across 7 conditions for the SCI rower. All able-bodied participants successfully rowed at required knee ranges of motion and speeds. The SCI rower managed to row under 4 out of 6 conditions with acceptable variations from the desired form (Table 2) and had difficulty returning to the most forward position for the other two conditions (45° – 135°, 40 RPM and 45° – 165°, 25 RPM). The SCI rower’s self-selected form resulted in 0.39 BW foot reaction force and 1.83 BW peak compressive tibiofemoral force.

**Table 2.**
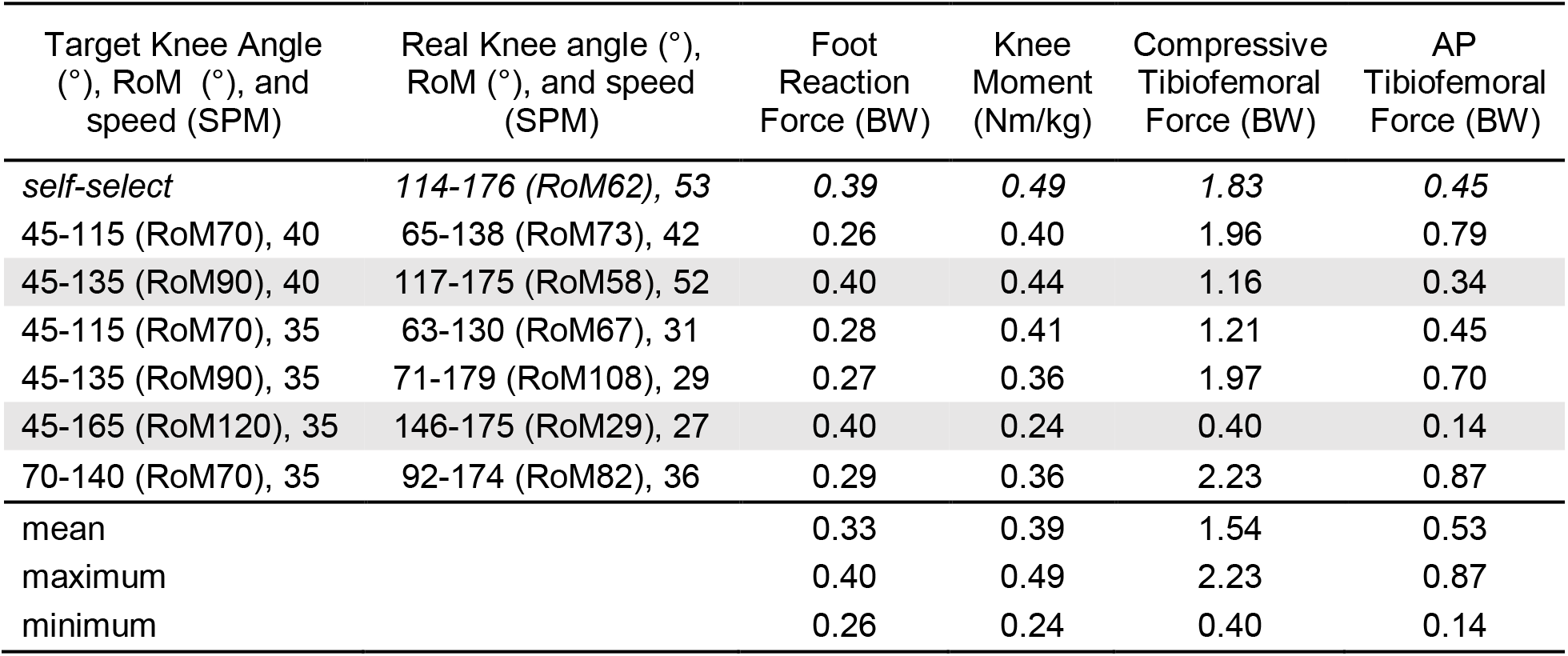
Normalized peak foot reaction force, peak knee extension moment, peak compressive tibiofemoral force, and peak anterior-posteior (AP) tibiofemoral force under self-selected rowing style and 6 testing conditions in FES-rowing from a spinal cord injured (SCI) participant. Shaded area indicates trials that the participant was unable to meet the target. RoM: ranges of motion, SPM: strokes per minute.

| Target Knee Angle (°), RoM (°), and speed (SPM) | Real Knee angle (°), RoM (°), and speed (SPM) | Foot Reaction Force (BW) | Knee Moment (Nm/kg) | Compressive Tibiofemoral Force (BW) | AP Tibiofemoral Force (BW) |
| --- | --- | --- | --- | --- | --- |
| *self-select* | 114-176 (RoM62), 53 | 0.39 | 0.49 | 1.83 | 0.45 |
| 45-115 (RoM70), 40 | 65-138 (RoM73), 42 | 0.26 | 0.40 | 1.96 | 0.79 |
| 45-135 (RoM90), 40 | 117-175 (RoM58), 52 | 0.40 | 0.44 | 1.16 | 0.34 |
| 45-115 (RoM70), 35 | 63-130 (RoM67), 31 | 0.28 | 0.41 | 1.21 | 0.45 |
| 45-135 (RoM90), 35 | 71-179 (RoM108), 29 | 0.27 | 0.36 | 1.97 | 0.70 |
| 45-165 (RoM120), 35 | 146-175 (RoM29), 27 | 0.40 | 0.24 | 0.40 | 0.14 |
| 70-140 (RoM70), 35 | 92-174 (RoM82), 36 | 0.29 | 0.36 | 2.23 | 0.87 |
| mean |  | 0.33 | 0.39 | 1.54 | 0.53 |
| maximum |  | 0.40 | 0.49 | 2.23 | 0.87 |
| minimum |  | 0.26 | 0.24 | 0.40 | 0.14 |

### Effect of rowing speed

There was a significant speed effect on peak foot reaction force (p < 0.001) and peak knee extension moment in able-bodied rowers (p < 0.001); faster speed resulted in greater foot reaction force and knee moment at both 70° and 90° knee RoM. Compared to 25 SPM, rowing at 40 SPM initiated 0.16 BW greater foot reaction force (p < 0.001) and 0.35 Nm/kg higher knee moment (p < 0.001), and rowing at 35 SPM resulted in 0.11 BW greater foot reaction force (p < 0.001) and 0.27 Nm/kg higher knee moment (p < 0.001), with 70° knee RoM. Compared to 35 SPM, rowing at 40 SPM initiated 0.05 BW greater foot reaction force (p < 0.001) and 0.08 Nm/kg higher knee moment (p = 0.037) with 70° knee RoM. When rowing with 90° knee RoM, foot reaction force (p < 0.001) and knee moment (p < 0.001) were significantly different between 25 SPM and 35 SPM and between 25 SPM and 40 SPM conditions (Figure 1).

**Figure 1.**
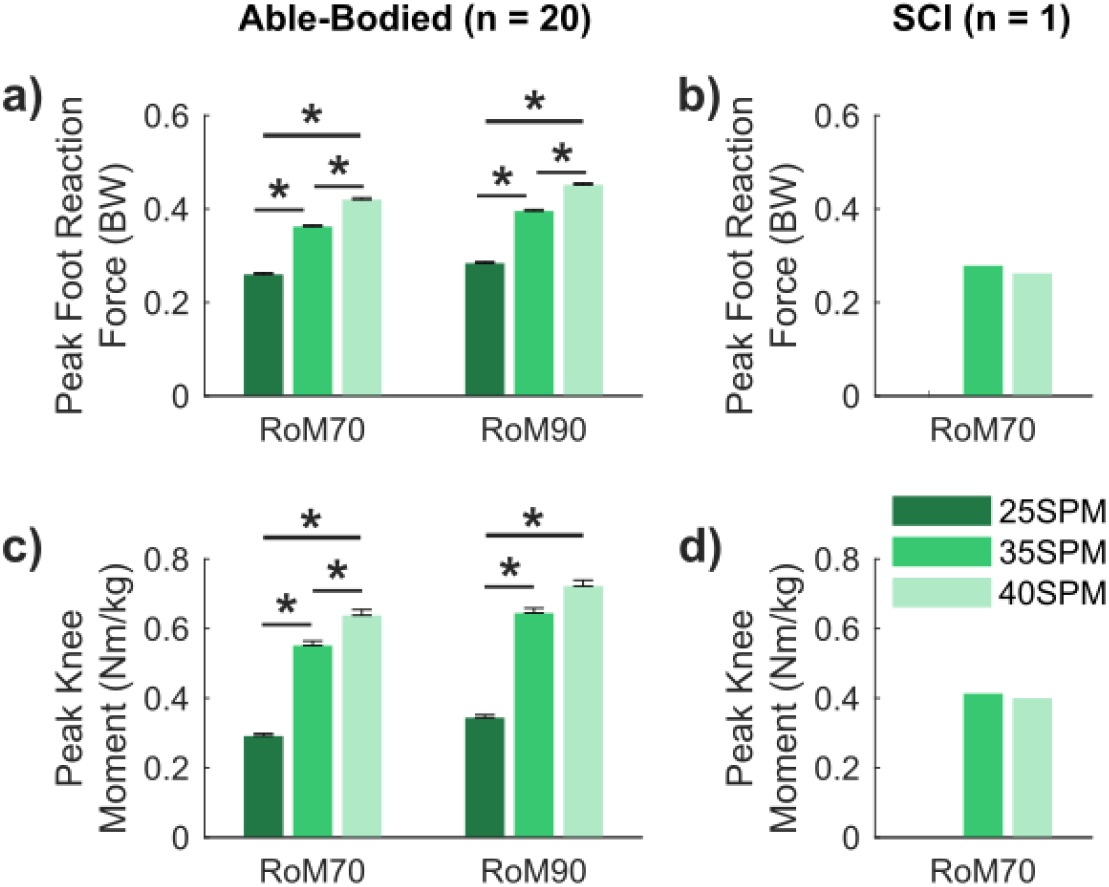
a) Normalized peak foot reaction force and c) normalized peak knee extension moment of 20 able-bodied participants rowing at three different speeds with knee ranges of motion (RoM) of 70° and 90°. Error bars indicate standard errors, and * indicates significant differences with p < 0.05. b) normalized peak foot reaction force and d) normalized peak knee extension moment of one spinal cord injured (SCI) participant rowing at two speeds (35 SPM and 40 SPM) with 70° knee RoM.

For the SCI rower, peak foot reaction force decreased by 0.02 BW, peak knee moment decreased by 0.01 Nm/kg, and peak compressive tibiofemoral force increased by 0.75 BW when speed increased from 35 SPM (actual speed: 31 SPM) to 40 SPM (actual speed: 42 SPM) with 70° knee RoM (Figure 4, Table 2).

### Effect of knee RoM

Knee RoM had a significant effect on peak foot reaction force (p = 0.003) and peak knee extension moment (p = 0.012) in able-bodied participants. Larger knee RoM resulted in greater foot reaction force and knee moment at both 25 SPM and 35 SPM. Regardless of rowing speed (25 SPM or 35 SPM), foot reaction force (p ≤ 0.009) and knee moment (p ≤ 0.037) were significantly lower during rowing with 70° compared to 90° or 120° RoM conditions (Figure 2).

**Figure 2.**
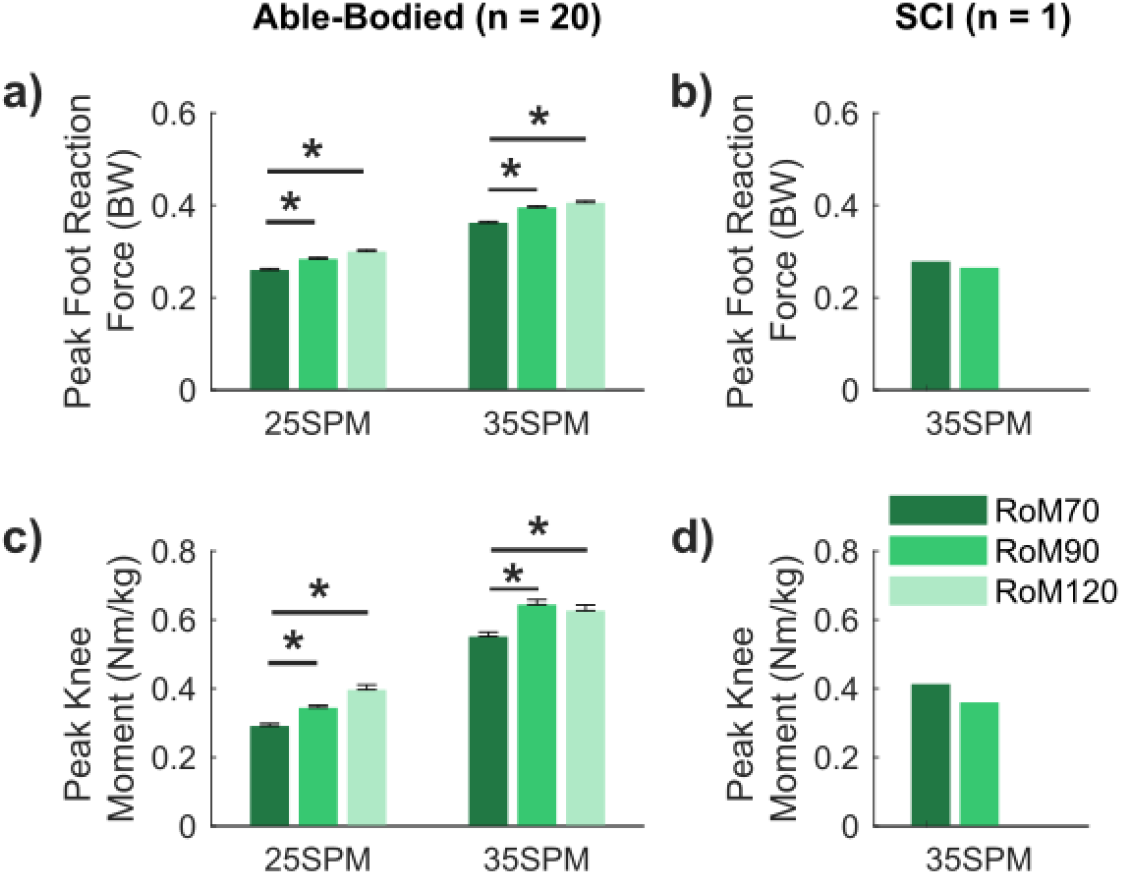
a) Normalized peak foot reaction force and c) normalized peak knee extension moment of 20 able-bodied participants rowing at three different knee ranges of motion (RoM) at 25 and 35 strokes per minute (SPM). Error bars indicate standard errors, and * indicates significant differences with p < 0.05. b) normalized peak foot reaction force and d) normalized peak knee extension moment of one spinal cord injured (SCI) participant rowing with two knee RoM (70° and 90°) at 35 SPM.

The SCI rower had 0.01 BW smaller peak foot reaction force, 0.05 Nm/kg smaller peak knee moment, and 0.76 BW greater peak compressive tibiofemoral force when rowing with 90° compared to 70° knee RoM at 35 SPM (actual speed: 29 SPM) (Figure 4, Table 2).

### Effect of seat position

In able-bodied rowers, seat position had a significant effect on peak foot reaction force (p < 0.001) and peak knee extension moment (p < 0.001). A more rear seat position resulted in smaller foot reaction force and knee moment at both 25 SPM and 35 SPM. Compared to a forward seat position, rowing with a rear seat position significantly decreased peak foot reaction force (p ≤ 0.001) and peak knee moment (p ≤ 0.004) at both 25 SPM and 35 SPM rowing speeds. Rowing with a middle seat position resulted in significantly decreased peak knee moment (p ≤ 0.007) and similar peak foot reaction force compared to a forward seat position at both 25 SPM and 35 SPM (Figure 3).

**Figure 3.**
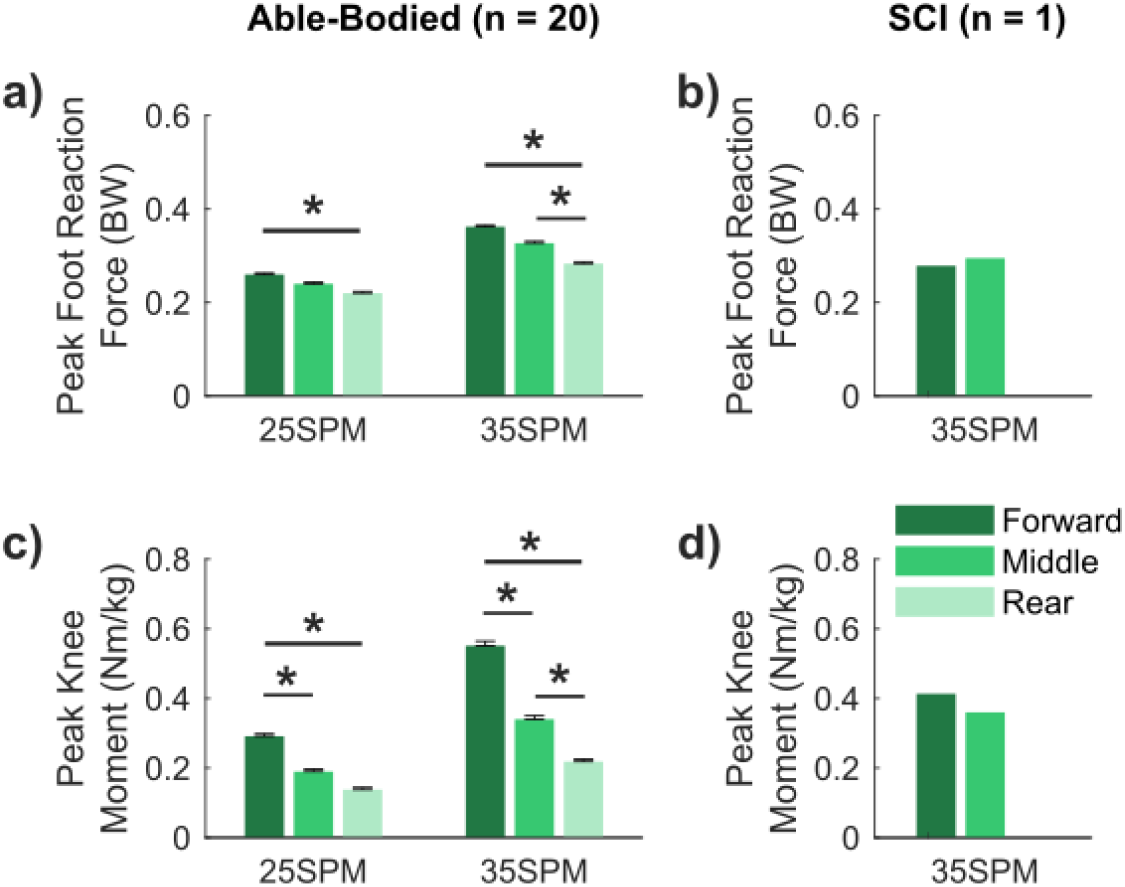
a) Normalized peak foot reaction force and c) normalized peak knee extension moment of 20 able-bodied participants rowing at three seat positions at 25 and 35 strokes per minute (SPM). Error bars indicate standard errors, and * indicates significant differences with p < 0.05. b) normalized peak foot reaction force and d) normalized peak knee extension moment of one spinal cord injured (SCI) participant rowing with two seat positions (Forward and Middle) at 35 SPM.

**Figure 4.**
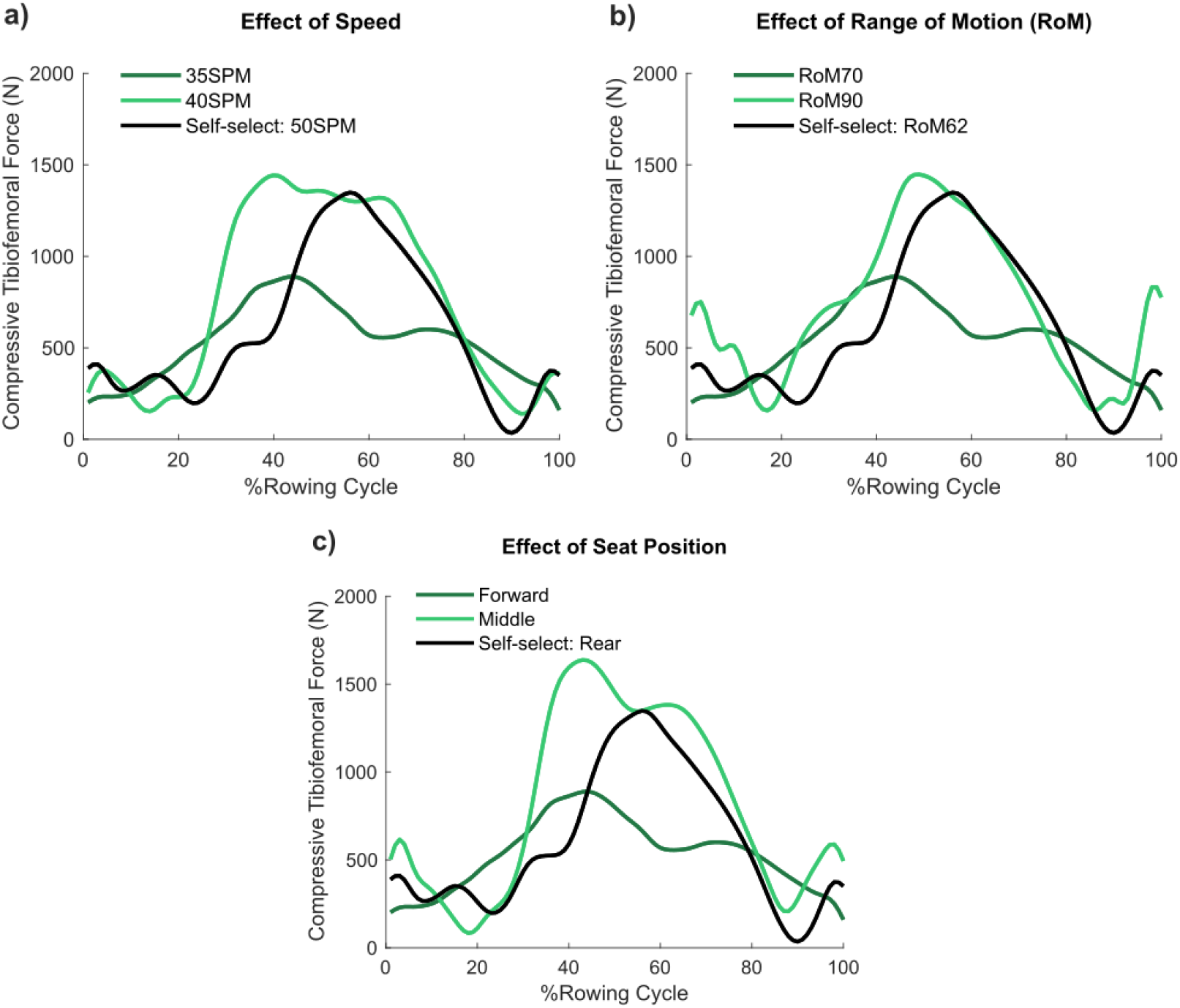
Compressive tibiofemoral force of a rowing cycle when the spinal cord injured (SCI) participant rowed using self-selected style and a) at two speeds, b) with two knee ranges of motion (RoM), and c) with two seat positions.

The SCI rower had 0.01 BW greater peak foot reaction force, 0.05 Nm/kg smaller knee moment, and 1.02 BW greater peak compressive tibiofemoral force while rowing with a middle compared to a forward seat position at 35 SPM (Figure 4, Table 2).

## Discussion

We investigated the effects of adapted rowing ergometer setup and speed on peak foot reaction force and knee extension moment in a group of able-bodied adults and a single SCI subject. The hypotheses were supported that rowing speed, knee RoM, and seat position all affected lower extremity loading in able-bodied and SCI rowers. In able-bodied rowers, having a forward starting position for the seat, combined with a fast speed and large knee RoM, was associated with the greatest foot reaction force and knee moment. However, this combination did not yield the greatest foot reaction force for the SCI rower, who generated a greater foot reaction force by rowing with a smaller knee RoM and a more extended knee.

The able-bodied and SCI participants generated lower foot reaction forces than what has been previously reported. When novice able-bodied individuals rowed on a normal ergometer, the foot reaction force ranged from 0.53 to 0.98 BW^27,28^. Using the adapted ergometer in this study, all participants generated only half the foot reaction force: 0.28 to 0.45 BW for the able-bodied and 0.26 to 0.40 BW for the SCI rower. The adaptations made to the ergometer changed the rowing biomechanics. With a normal ergometer, larger foot reaction forces can be achieved by increasing trunk extension at the end of the drive phase, ankle flexion at the beginning of the drive phase, and having a larger hip RoM^16^. On an adapted ergometer, the added seat back and seatbelt constrain both trunk and hip movement, and the front and rear stoppers limit ankle motion. These changes also limited torque generation at the joint: peak knee moment was 0.64 Nm/kg among our participants, compared to 0.89 Nm/kg in novice rowers who rowed at the same speed on normal ergometers^27^.

Rowing speed and ergometer setup had a significant influence on foot force production in both able-bodied and SCI rowers. The able-bodied participants generated twice as much foot reaction force when rowing with a fast speed (40 SPM), large knee RoM (90° or 120°), and a forward seat position compared to other conditions. Higher rowing speed causes greater acceleration and larger knee RoM allows joints to generate more power, both of which contributed to higher force at the foot. Larger knee RoM or faster speed, however, did not yield higher foot force or knee moment in the SCI rower. This highlights the inherent difference in the behavior of electrically stimulated muscles and voluntarily contracted muscles. The FES stimulator delivers a fixed amount of energy to the muscle, which is either used to generate motion or can be transmitted through segments until it reaches the foot stretcher, reflecting as the foot reaction force. In other words, the foot reaction force could be compromised if a larger range of motion is to be completed. This does not apply to able-bodied rowers, who can adjust muscle recruitment depending on the task. For example, they can engage additional muscles or motor units within a given muscle to complete a trial that requires higher energy (i.e., rowing with a large knee RoM or faster speed) and generate a larger foot reaction force.

The effects of seat position on foot reaction force were also different between able-bodied rowers and the SCI rower: the former generated higher force at a forward seat position and the latter at a more rear position. This indicates that the optimum joint working range may exist and be different for electrically-stimulated muscles and voluntarily contracted muscles.

The SCI rower generated a relatively large force and moment in self-selected style, with the force magnitude (0.36 BW) reaching the upper limit of previously reported foot reaction forces in SCI rowers (0.15 – 0.35 BW)^12,14,29^. This appeared to result from a combination of the rower set-up, good hand-leg coordination, and an experienced and strong participant. The individual in the current study had rowed for six years, which made him more experienced than those included in previous studies. He also began rowing very shortly after becoming spinal cord injured and had no obvious leg muscle atrophy compared to many individuals with chronic SCI.

A common goal of FES-rowing for individuals with SCI is to prevent bone loss by loading the lower extremities. FES during active-resisted standing generated 1.50 BW knee contact force in individuals with SCI, which significantly attenuated bone loss at the tibia and femur ^30^. The individual with SCI in the present study received 0.40 to 2.23 BW compressive tibiofemoral force when rowing in various forms, suggesting that ergometer setup and speed are crucial factors in determining whether the exercise will be beneficial to bone. Furthermore, the large variance in reported knee joint loading (1.25 – 4.6 BW) in previous FES-rowing studies could be partially attributed to the difference in rowing forms and setups between subjects^13,29^. The results show that the external loading (foot reaction force) was much smaller than internal knee joint loading, suggesting that muscle contraction was the primary contributor to tibiofemoral force in FES-rowing. Therefore, applying FES at specific knee ranges of motion could optimize muscle force generation and consequently maximize knee loading. The findings bring up an issue that warrants further researchers to investigate with more SCI rowers.

One limitation of the current study is that only the right foot reaction force was measured. We argue that rowing is symmetrical, especially when both legs are constrained by the knee stabilizer. A previous study that measured bilateral foot reaction forces in FES-rowing reported a 7% difference between the left and right foot reaction force. Here, the kinematic data showed nearly identical joint angles for the left and right legs. Another limitation is that we only tested on one participant with SCI. Although this participant does not represent all individuals with SCI, the data are important to inform future investigations on FES-rowing biomechanics. In particular, this case study highlights that there may be a trade-off between producing force and producing motion in FES-rowing due to the inability to recruit additional motor units and/or muscles in SCI users. In our musculoskeletal model of the SCI participant, we limited the number of muscles that could be activated and did not consider any possibility of muscle spasm. Thus far, there is no standard way to measure muscle spasms, nor is there a muscle model to takes this factor into account. Future studies could validate muscle activation with EMG measurements. Our validation compared the onset/offset times predicted by the musculoskeletal model to when the stimulator was triggered by the user and showed good agreement (The quadriceps was activated 53 ms after the subject pressed the button, and the hamstrings was activated 45 ms before the subject completely released the button.). The consistency of these results gives us confidence in the model predictions.

## Conclusions

The design of the adapted ergometer, which constrained trunk motion, limited the amount of force that could be produced during rowing. Changing ergometer setup affected force production for all users, but the setup that yielded the highest foot reaction forces and knee moments was different for able-bodied versus SCI rowers. Able-bodied rowers adopted a fast rowing speed, forward seat position, and large knee RoM. In contrast, the SCI rower had a rear seat position and smaller knee RoM. Our findings support future research with a larger cohort with SCI rowers, and suggest rowing setups and forms to be considered during FES-rowing at home or in clinic settings.

## Data Availability

There is no supplementary material.

## References

1. Tan, C. O., Battaglino, R. A. & Morse, L. R. Spinal Cord Injury and Osteoporosis: Causes, Mechanisms, and Rehabilitation Strategies. Int. J. Phys. Med. Rehabil. 1, 127 (2013).

2. Panisset, M. G., Galea, M. P. & El-Ansary, D. Does early exercise attenuate muscle atrophy or bone loss after spinal cord injury? Spinal Cord (2016) doi:10.1038/sc.2015.150.

3. Hammond, E. R., Metcalf, H. M., McDonald, J. W. & Sadowsky, C. L. Bone Mass in Individuals With Chronic Spinal Cord Injury: Associations With Activity-Based Therapy, Neurologic and Functional Status, a Retrospective Study. Arch. Phys. Med. Rehabil. 95, 2342–2349 (2014).

4. Dolbow, D. R. et al. The effects of spinal cord injury and exercise on bone mass: A literature review. NeuroRehabilitation (2011) doi:10.3233/NRE-2011-0702.

5. Chen, S.-C. et al. Increases in bone mineral density after functional electrical stimulation cycling exercises in spinal cord injured patients. Disabil. Rehabil. 27, 1337–1341 (2005).

6. Dudley-Javoroski, S. et al. High dose compressive loads attenuate bone mineral loss in humans with spinal cord injury. Osteoporos. Int. (2012) doi:10.1007/s00198-011-1879-4.

7. Groah, S. L., Lichy, A. M., Libin, A. V. & Ljungberg, I. Intensive Electrical Stimulation Attenuates Femoral Bone Loss in Acute Spinal Cord Injury. PM R (2010) doi:10.1016/j.pmrj.2010.08.003.

8. Andrews, B. et al. A Design Method for FES Bone Health Therapy in SCI. Eur. J. Transl. Myol. 26, (2016).

9. Morse, L. R. et al. Combination Therapy With Zoledronic Acid and FES-Row Training Mitigates Bone Loss in Paralyzed Legs: Results of a Randomized Comparative Clinical Trial. JBMR Plus 3, e10167 (2019).

10. Lambach, R. L. et al. Bone changes in the lower limbs from participation in an FES rowing exercise program implemented within two years after traumatic spinal cord injury. Journal of Spinal Cord Medicine (2018) doi:10.1080/10790268.2018.1544879.

11. Deley, G., Denuziller, J., Casillas, J. M. & Babault, N. One year of training with FES has impressive beneficial effects in a 36-year-old woman with spinal cord injury. J. Spinal Cord Med. (2017) doi:10.1080/10790268.2015.1117192.

12. Draghici, A. E., Picard, G., Taylor, J. A. & Shefelbine, S. J. Assessing kinematics and kinetics of functional electrical stimulation rowing. J. Biomech. 53, 120–126 (2017).

13. Gibbons, R. S. et al. Can FES-rowing mediate bone mineral density in SCI: a pilot study. Spinal Cord 52, S4–S5 (2014).

14. Halliday, S. E., Zavatsky, A. B. & Hase, K. Can functional electric stimulation-assisted rowing reproduce a race-winning rowing stroke? Arch. Phys. Med. Rehabil. (2004) doi:10.1016/j.apmr.2003.11.025.

15. Černe, T., Kamnik, R., Vesnicer, B., Žganec Gros, J. & Munih, M. Differences between elite, junior and non-rowers in kinematic and kinetic parameters during ergometer rowing. Hum. Mov. Sci. (2013) doi:10.1016/j.humov.2012.11.006.

16. Buckeridge, E. M., Bull, A. M. J. & McGregor, A. H. Biomechanical determinants of elite rowing technique and performance. Scand. J. Med. Sci. Sports 25, e176–e183 (2015).

17. Vinther, A. et al. Slide-based ergometer rowing: Effects on force production and neuromuscular activity. Scand. J. Med. Sci. Sport. (2013) doi:10.1111/j.1600-0838.2011.01441.x.

18. Millar, S. et al. Elite Rowers Apply Different Forces Between Stationary and Sliding Ergometers, & on-Water Rowing. in Biomechanics in Sports (2017).

19. Buckeridge, E. M., Weinert-Aplin, R. A., Bull, A. M. J. & McGregor, A. H. Influence of foot-stretcher height on rowing technique and performance. Sport. Biomech. (2016) doi:10.1080/14763141.2016.1185459.

20. Taylor, J. A., Picard, G. & Widrick, J. J. Aerobic Capacity With Hybrid FES Rowing in Spinal Cord Injury: Comparison With Arms-Only Exercise and Preliminary Findings With Regular Training. PM R (2011) doi:10.1016/j.pmrj.2011.03.020.

21. Wu, G. et al. ISB recommendation on definitions of joint coordinate system of various joints for the reporting of human joint motion - Part I: Ankle, hip, and spine. Journal of Biomechanics (2002) doi:10.1016/S0021-9290(01)00222-6.

22. Winter, D. A. Moments of force and mechanical power in jogging. J. Biomech. 16, 91–97 (1983).

23. Delp, S. L. et al. OpenSim: Open-source software to create and analyze dynamic simulations of movement. IEEE Trans. Biomed. Eng. (2007) doi:10.1109/TBME.2007.901024.

24. Lai, A. K. M., Arnold, A. S. & Wakeling, J. M. Why are Antagonist Muscles Co-activated in My Simulation? A Musculoskeletal Model for Analysing Human Locomotor Tasks. Ann. Biomed. Eng. (2017) doi:10.1007/s10439-017-1920-7.

25. Anderson, F. C. & Pandy, M. G. Static and dynamic optimization solutions for gait are practically equivalent. J. Biomech. (2001) doi:10.1016/S0021-9290(00)00155-X.

26. Fang, Y., Morse, L. R., Nguyen, N., Tsantes, N. G. & Troy, K. L. Anthropometric and biomechanical characteristics of body segments in persons with spinal cord injury. J. Biomech. (2017) doi:10.1016/j.jbiomech.2017.01.036.

27. Černe, T., Kamnik, R. & Munih, M. The measurement setup for real-time biomechanical analysis of rowing on an ergometer. Meas. J. Int. Meas. Confed. (2011) doi:10.1016/j.measurement.2011.09.006.

28. Hase, K., Kaya, M., Zavatsky, A. B. & Halliday, S. E. Musculoskeletal loads in ergometer rowing. J. Appl. Biomech. (2004) doi:10.1123/jab.20.3.317.

29. Chandran, V. D. et al. Tibiofemoral forces during FES rowing in individuals with spinal cord injury. Comput. Methods Biomech. Biomed. Engin. 1–14 (2020) doi:10.1080/10255842.2020.1821880.

30. Dudley-Javoroski, S. & Shields, R. K. Active-resisted stance modulates regional bone mineral density in humans with spinal cord injury. J. Spinal Cord Med. (2013) doi:10.1179/2045772313Y.0000000092.

